# Characterization of antibody response in asymptomatic and symptomatic SARS-CoV-2 infection

**DOI:** 10.1101/2021.03.29.21254534

**Authors:** S Marchi, S Viviani, EJ Remarque, A Ruello, E Bombardieri, V Bollati, GP Milani, A Manenti, G Lapini, A Rebuffat, E Montomoli, CM Trombetta

## Abstract

SARS-CoV-2 pandemic is causing high morbidity and mortality burden worldwide with unprecedented strain on health care systems.

To elucidate the mechanism of infection, protection, or rapid evolution until fatal outcome of the disease we performed a study in hospitalized COVID-19 patients to investigate the time course of the antibody response in relation to the outcome. In comparison we investigated the time course of the antibody response in SARS-CoV-2 asymptomatic subjects.

Study results show that patients produce a strong antibody response to SARS-CoV-2 with high correlation between different viral antigens (spike protein and nucleoprotein) and among antibody classes (IgA, IgG, and IgM and neutralizing antibodies). The peak is reached by 3 weeks from hospital admission followed by a sharp decrease. No difference was observed in any parameter of the antibody classes, including neutralizing antibodies, between subjects who recovered or with fatal outcome. Only few asymptomatic subjects developed antibodies at detectable levels.

## INTRODUCTION

On March 11, 2020, the World Health Organization (WHO) Director General declared a pandemic situation due to a novel coronavirus causing a Severe Acute Respiratory Syndrome (SARS) rapidly spreading worldwide [1]. The novel coronavirus (CoV) SARS-CoV-2 has been firstly identified in Wuhan, the capital city of Hubei Province, China, at the end of 2019 when a cluster of atypical pneumonia was identified [1, 2]. In January 2020, SARS-CoV-2 was isolated and sequenced as a CoV genetically related to the highly pathogenic CoV (SARS-CoV-1) responsible for the 2003 SARS outbreak. CoV s are known to cause disease in humans. There are four CoVs responsible for the common cold and another one highly pathogenic known as Middle Eastern Respiratory Syndrome (MERS)-CoV [2-6]. SARS-CoV-1 caused a global pandemic with approximately 10% case fatality rate (CFR) [7]. However, SARS-CoV-1 has stopped circulating in humans since 2004. MERS-CoV was first reported from Saudi Arabia in 2012 and has continued to infect humans in 27 countries with limited human-to-human transmission, and a CFR of approximately 34.4%, according to the most recent WHO report [7]. As SARS-CoV-1 and MERS-CoV, the SARS-CoV-2 virus is an enveloped, single-stranded, and positive-sense RNA virus belonging to the Betacoronavirus Genus, *Coronaviridae* family. The genome of SARS-CoV-2, as the other emerging pathogenic human CoVs, encodes four major structural proteins: spike (S), envelope (E), membrane (M), nucleocapsid (N); approximately 16 nonstructural proteins (nsp1–16), and five to eight accessory proteins. Among them, the S protein plays an essential role in viral attachment, fusion, entry, and transmission. The S protein is the common target antigen for neutralizing antibodies and vaccine development [8-11]. SARS-CoV-2 predominant transmission way was identified early to be a human-to-human mode occurring through respiratory droplets, however, close contact with infected surfaces or objects may also be an occasional way of transmission. The virus is excreted and detectable in saliva and stool [5, 9, 12]. SARS-CoV-2 disease, or COVID-19, ranges from a mild upper/lower respiratory tract infection that resolves in a few days without sequelae to more serious disease with fever, cough, shortness of breath, myalgias, fatigue, confusion, headache, sore throat, acute respiratory distress syndrome, leading to respiratory or multiorgan failure [5, 9, 12]. The fatality rate is high in people with underlying comorbidities, particularly in the elderly, such as diabetes, hypertension, chronic respiratory disease or cardiovascular disease [9, 12]. After almost one year after the first cases were reported in Wuhan, as of 13 December 2020, there have been over 70 million cases and over 1.5 million deaths reported to WHO of confirmed COVID-19 [13] with Europe being one of the most affected areas, second only to the Americas. COVID-19 pandemic is causing high morbidity and mortality burden worldwide, an unprecedented strain on health care systems, with social and economic disruption as the only effective prevention is social distancing leading entire countries in quarantine for weeks or months with dramatic impact on day-to-day human, social and economic life [14].

Italy has been one of the earliest and most affected countries by COVID-19. Although the first SARS-CoV-2 case identified in Codogno at the end of February 2020 is considered the Italian index case, some evidence has later arisen that the virus had circulated in Italy and Europe since autumn 2019 [15-18]. Italy suffered the first wave from February until June 2020 when the whole country was under strict lockdown. During this period the most affected areas were located in Northern and to a less extent in Central Italy, while the Southern part of the country was relatively unaffected [19, 20]. During the summer period until the end of September 2020 COVID-19 remained endemic, with a second epidemic wave starting in October 2020 that led to a second national lockdown in November 2020. As of the 13^th^ of December 2020, more than 1.8 million confirmed cases and more than 64.000 deaths due to SARS-CoV-2 were reported to ISS (Istituto Superiore di Sanità), Rome [21]. The mean age of fatalities in COVID-19 infected people was 80 years, 42,4% were women and more than 90% had one or more co-morbidity as ischemic heart disease, diabetes, active cancer, atrial fibrillation, dementia, and a history of stroke [19].

The emerging and rapid diffusion of COVID-19 has risen the calls for more targeted research in the field [22] helping to elucidate the mechanism of infection, protection, or rapid evolution until fatal outcome of the disease. We present here a study performed in hospitalized infected COVID-19 patients to investigate the time course of the antibody response in relation to the outcome, and as explorative comparison, we also investigated the time course of the antibody response in SARS-CoV-2 asymptomatic subjects.

## MATERIAL AND METHODS

### Study population

This was a retrospective study on COVID-19 patients and SARS-CoV-2 asymptomatic subjects during the first epidemic wave occurred in Italy between March and May 2020.

A total of 42 COVID-19 patients, hospitalized at Humanitas Gavazzeni (Bergamo, Italy), were retrospectively selected for this study, of whom 35 (22 males and 13 females) recovered and 7 (3 males and 4 females) had a fatal outcome. All subjects were admitted to hospital with a diagnosis of interstitial pneumonia confirmed by chest radiograph or a CAT (computerized axial tomography) and had rhino-pharyngeal swab positive to SARS-CoV-2 (Real-Time PCR Thermo Fisher Scientific). Six patients required care in the intensive care unit (ICU), the others were hospitalized in the general medicine unit. Out of 7 deceased patients, 3 were hospitalized in ICU and 4 in the general medicine unit.

Serum samples were collected at different time points from March to April 2020 for diagnostic/therapeutic purposes. We selected patients who had available at least 5 blood samples during the period of hospital admission (baseline, day 2, day 6, day 12-14, day 18-20, day 27-30). Demographic and clinical variables reported in this study were those collected at hospital admission. For the purpose of this study patients were categorized according to the outcome: recovered or deceased.

During the first phase of the COVID-19 epidemic, little was known about this novel CoV and there was no standard therapy, so the management changed over the time. The Italian Society of Infectious and Tropical Diseases recommended as therapy hydroxychloroquine, antiviral agents, steroids, low molecular weight heparin and oxygen support in different combinations according to the clinician’s evaluation. The antibiotic therapy was adopted only in case of suspected or confirmed bacterial superinfection.

Serum samples from 25 asymptomatic subjects who presented a positive nasal swab for SARS-CoV-2 were collected as part of the UNICORN project and were analysed in the present study [23].

This study was approved by the Ethics Committee of the University of Siena (approval number 17373, approval date June 1, 2020), by the Ethics Committee of Humanitas Gavazzeni (approval number 236, approval date September 22, 2020 Protocol 670/20). The UNICORN study was approved by the Ethics Committee of the University of Milan (approval number 17/20, approval date March 6, 2020).

### Serological assay

#### ELISA

All serum samples were tested by commercial ELISA for the detection of IgA, IgG, and IgM against the S1 of SARS-CoV-2 (Aeskulisa^®^ SARS-CoV-2 S1 IgA, IgM, IgG, Aesku.Diagnostics, Wendelsheim, Germany) and for the detection of IgG against the nucleoprotein (NP) of SARS-CoV-2 (Aeskulisa^®^ SARS-CoV-2 NP IgG, Aesku.Diagnostics, Wendelsheim, Germany).

According to the manufacturer’s instructions, quantitative analysis was performed by use of a 4-parameter logistic standard curve obtained by plotting the optical density (OD) values measured for 4 calibrators against their antibody activity (U/ml) using logarithmic/linear coordinates. Antibody activities of the samples were evaluated from OD values using the generated curve and considered positive if >12 U/ml.

### Virus Neutralization assay

The virus neutralization (VN) assay has been performed as previously reported [24]. Briefly, serum samples were heat-inactivated for 30 minutes at 56°C and, starting from 1:10 dilution, were mixed with an equal volume of SARS-CoV-2 (2019 nCov/Italy INMI1 strain) viral solution containing 100 Tissue Culture Infective Dose 50% (TCID50). After 1 hour of incubation at room temperature, 100µl of virus-serum mixture were added to a 96-well plate containing an 80% confluent Vero E6 cell monolayer. Plates were incubated for 3 days at 37°C, 5% CO_2_ in humidified atmosphere, then inspected for presence/absence of cytopathic effect (CPE) by means of an inverted optical microscope. A CPE higher than 50% indicated infection. The VN titer was expressed as the reciprocal of the highest serum dilution showing protection from viral infection and CPE.

### Statistical analysis

All statistical analyses were performed using Microsoft R-Open version 3.5.0 (R Core Team (2018). R: A language and environment for statistical computing. R Foundation for Statistical Computing, Vienna, Austria. URL https://www.R-project.org/). For patient baseline characteristics continuous variables were evaluated using Mann-Whitney tests and for categorical variables Chi-square tests were used. Seroconversion rates were compared using Fisher’s exact test. Antibody levels were statistically evaluated using t-tests. Statistical significance was set at p<0.05, two tailed.

## RESULTS

### COVID-19 patients

Between March and April 2020, a total of 42 subjects were retrospectively selected for this study, of whom 35 recovered and 7 had a fatal outcome. The median age at admission was 64.0 years (interquartile range (IQR) 56.0-71.5) for those who recovered and 69.0 years (IQR 64.5-72.0) for deceased patients. The median length of stay in the hospital was similar in both groups with 11.0 days (IQR 9.0-24.5) and 10.0 (IQR 6.0-15.59) for recovered and deceased patients, respectively. The mean number of pre-existing conditions in recovered and deceased was 1.53 (standard deviation (SD) 1.25) and 2.0 (SD 1.41), respectively, and comorbidities were indicated in 1.88 (SD 1.36) and 3.5 (SD 3.54) of recovered and deceased patients, respectively. No differences were found between recovered and deceased patients when compared for symptoms at admission (fever, cough, diarrhea, dyspnea) or presence/absence of comorbidities and/or preexisting conditions. The other demographic, clinical, and blood chemistry variables collected at baseline were similar between the two groups, with exception of ALT that showed to be statistically significantly higher (p-value 0.021) in subjects who recovered (Table 1a).

**Table 1a.**
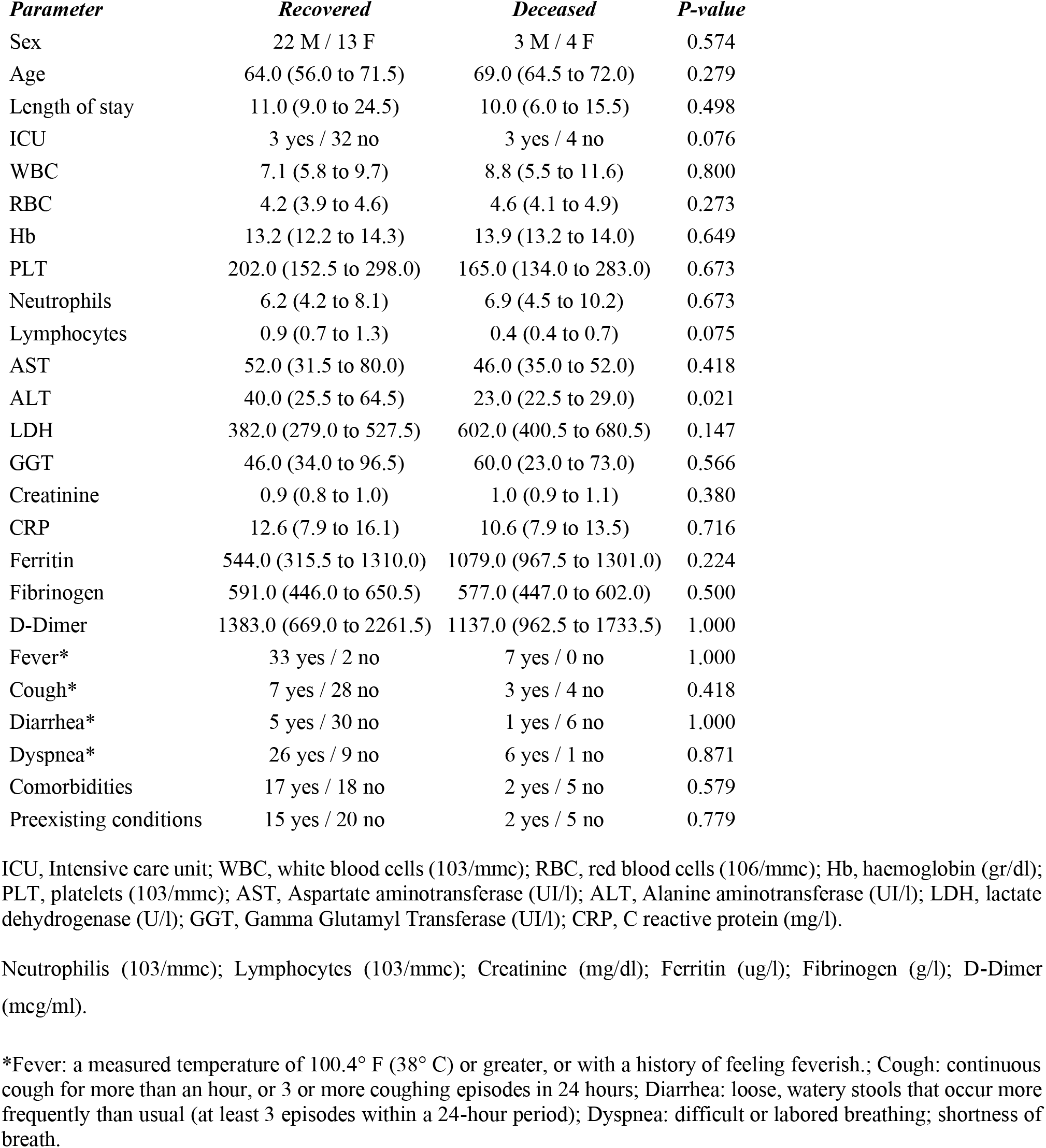
Baseline characteristics of COVID-19 patients according to outcome. Median (IQR)

At hospital admission, 15 subjects (35.7%) were negative for S1 IgM, 11 (26.2%) for S1 IgA, 13 (30.9%) for S1 IgG, 15 (35.7%) for NP IgG, and 10 (23.8%) for neutralizing antibodies. Five subjects (11.9%) were negative to any antibody assay at the time of admission; of these, 2 died and 3 recovered. Two days after admission, 6 subjects (14.3%) were still negative for S1 IgM, 7 (16.7%) for S1 IgA, 4 (9.5%) for S1 IgG, 3 (7.1%) for NP IgG, and 5 (11.9%) for neutralizing antibodies. Two subjects (4.8%) were still negative to any antibody assay; of these, 1 died and 1 recovered. At 6 days of sample collection, all subjects except one (97.6%) were positive to all assays (Figures 1-5). The exception was represented by a 40-year-old male subject, showing neither S1 IgM nor S1 IgA positivity in any sample, NP IgG with a borderline result at admission and at day 2, and negative at day 6, and neutralizing antibody titers less or equal than 40. This subject presented fever and dyspnea at admission with no comorbidities or preexisting conditions and recovered in 12 days.

**Figure 1.**
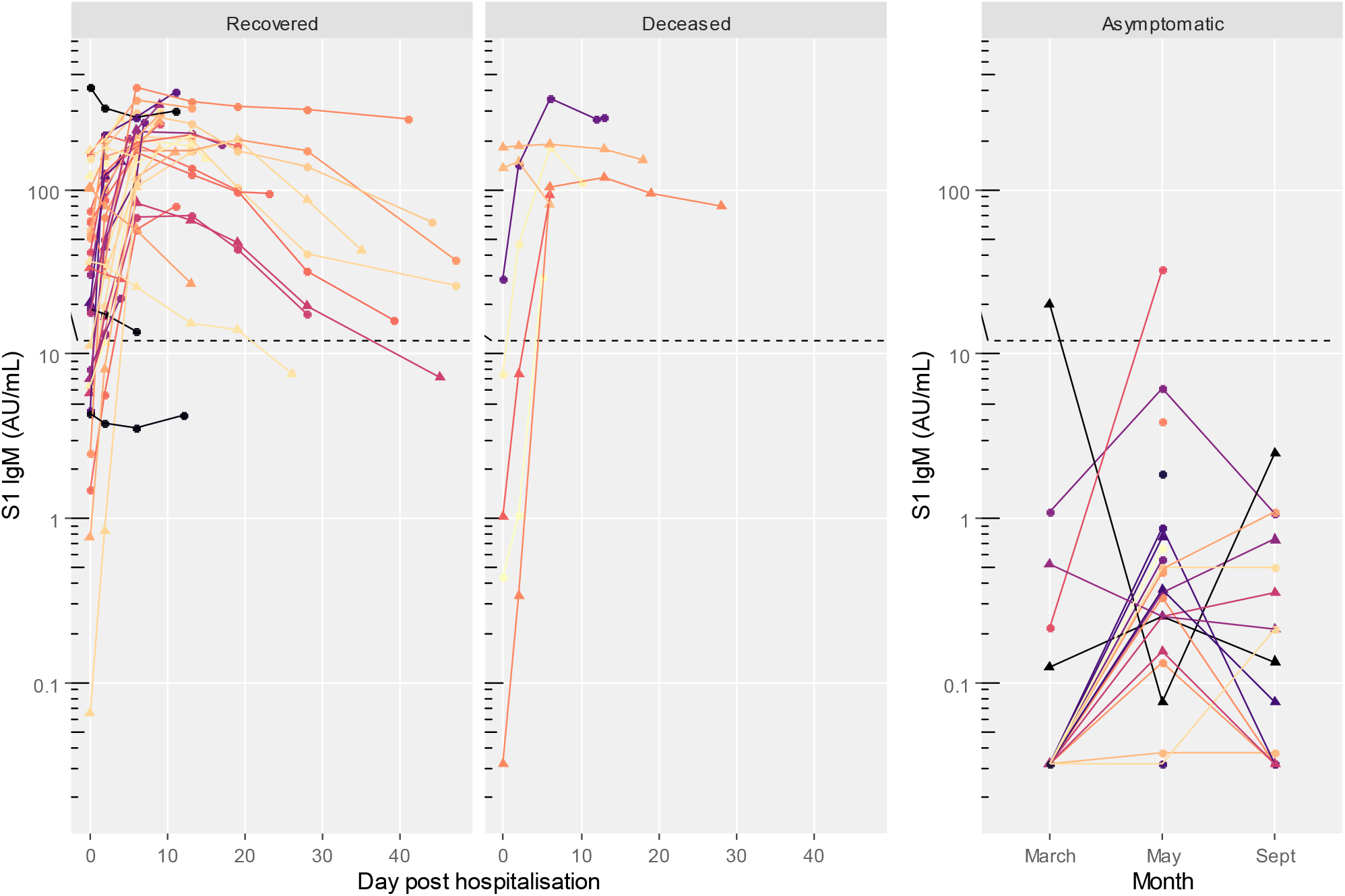
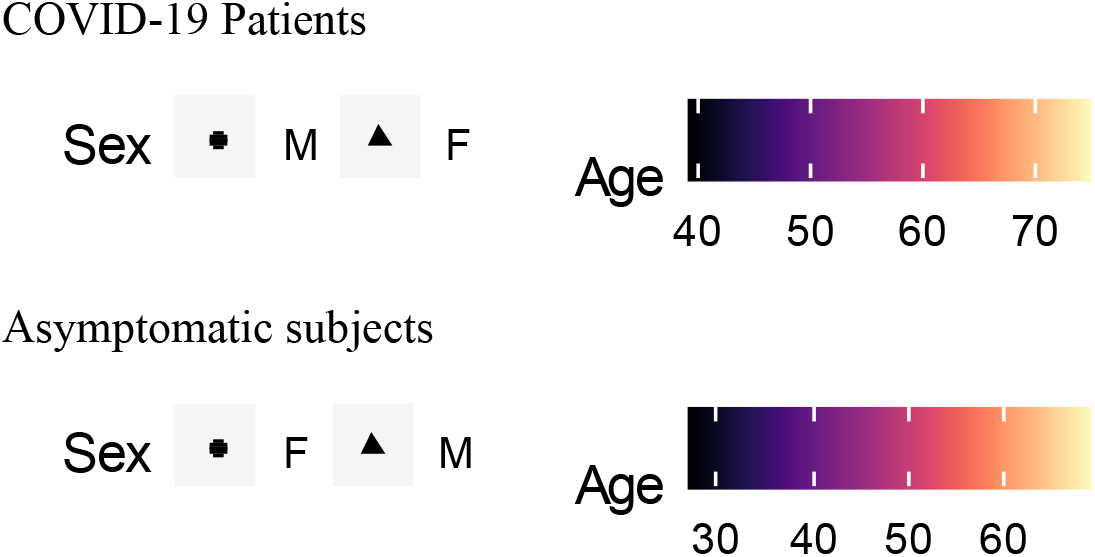
S1 IgM titres in COVID-19 patients (recovered and deceased) and asymptomatic subjects. Black dashed line indicates positivity threshold at 12 U/ml.

Two subjects, one recovered and one deceased both within 6 days after admission, were both negative to NP IgG at admission and at day 2. Two subjects, both recovered, were positive only for neutralizing antibodies at admission, with titers less or equal to 40. Both showed positivity to all antibodies tested at day 2 and later (Figures 1-5).

**Figure 2.**
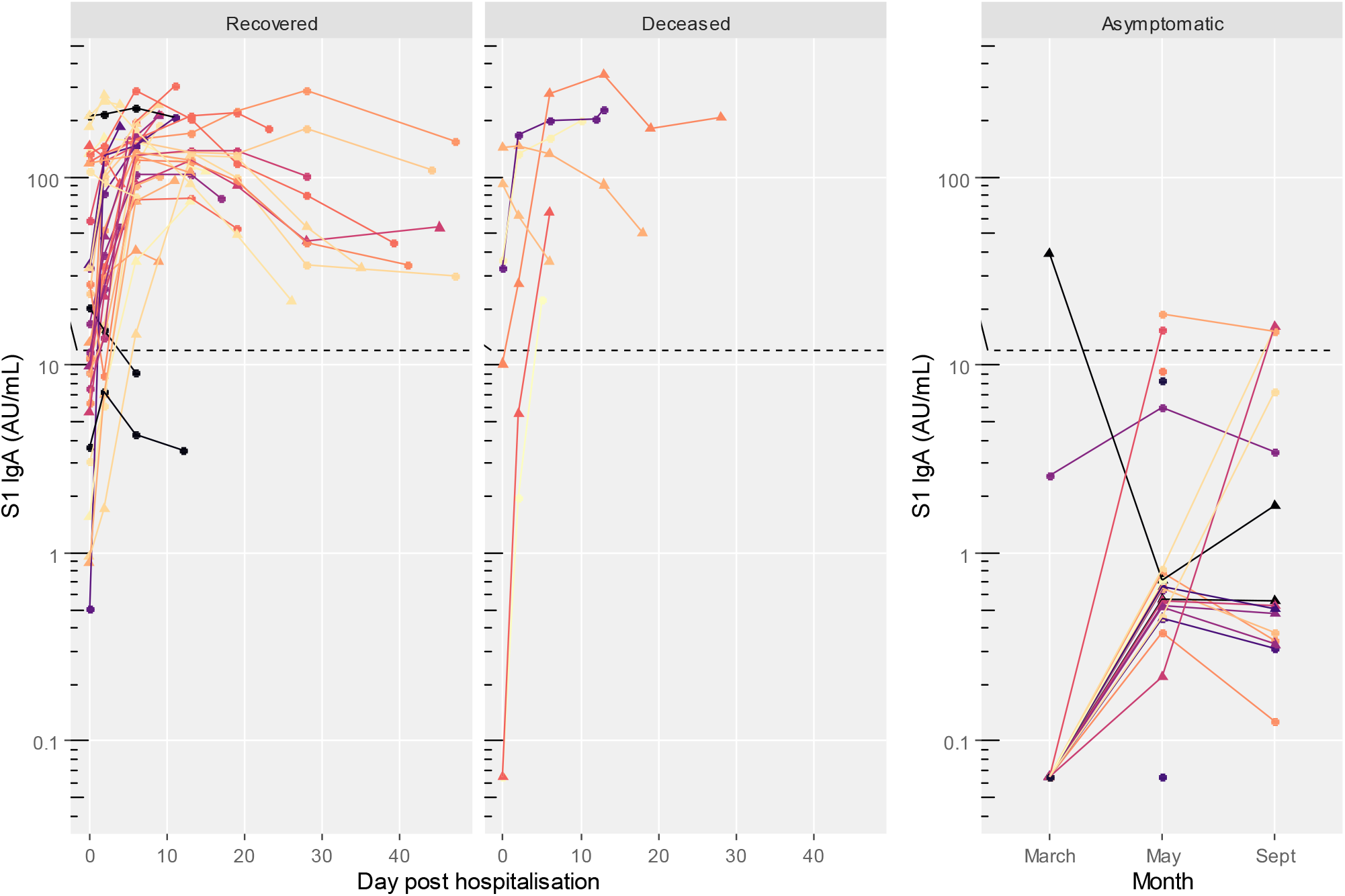
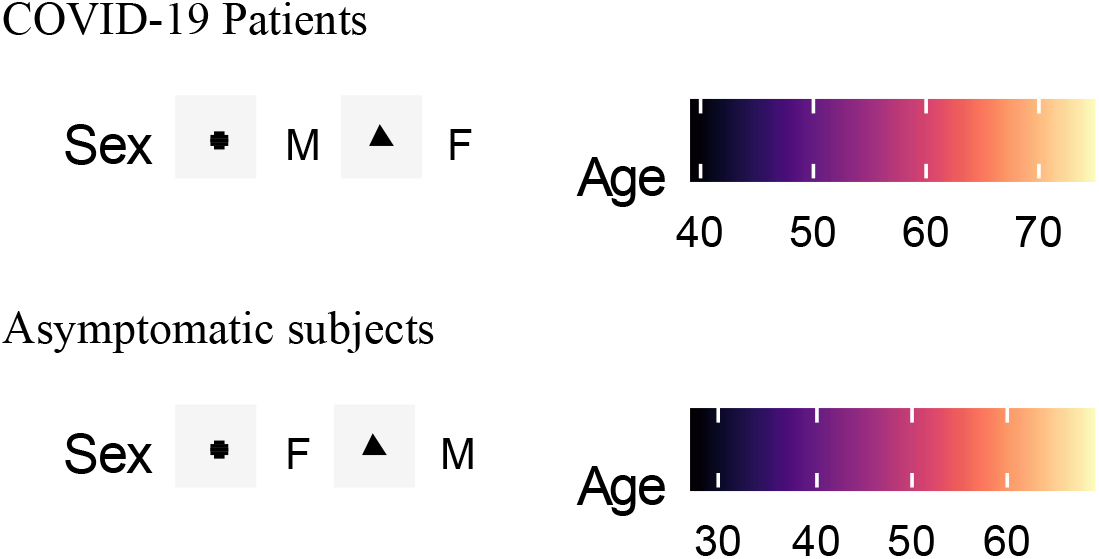
S1 IgA titres in COVID-19 patients (recovered and deceased) and asymptomatic subjects. Black dashed line indicates positivity threshold at 12 U/ml.

**Figure 3.**
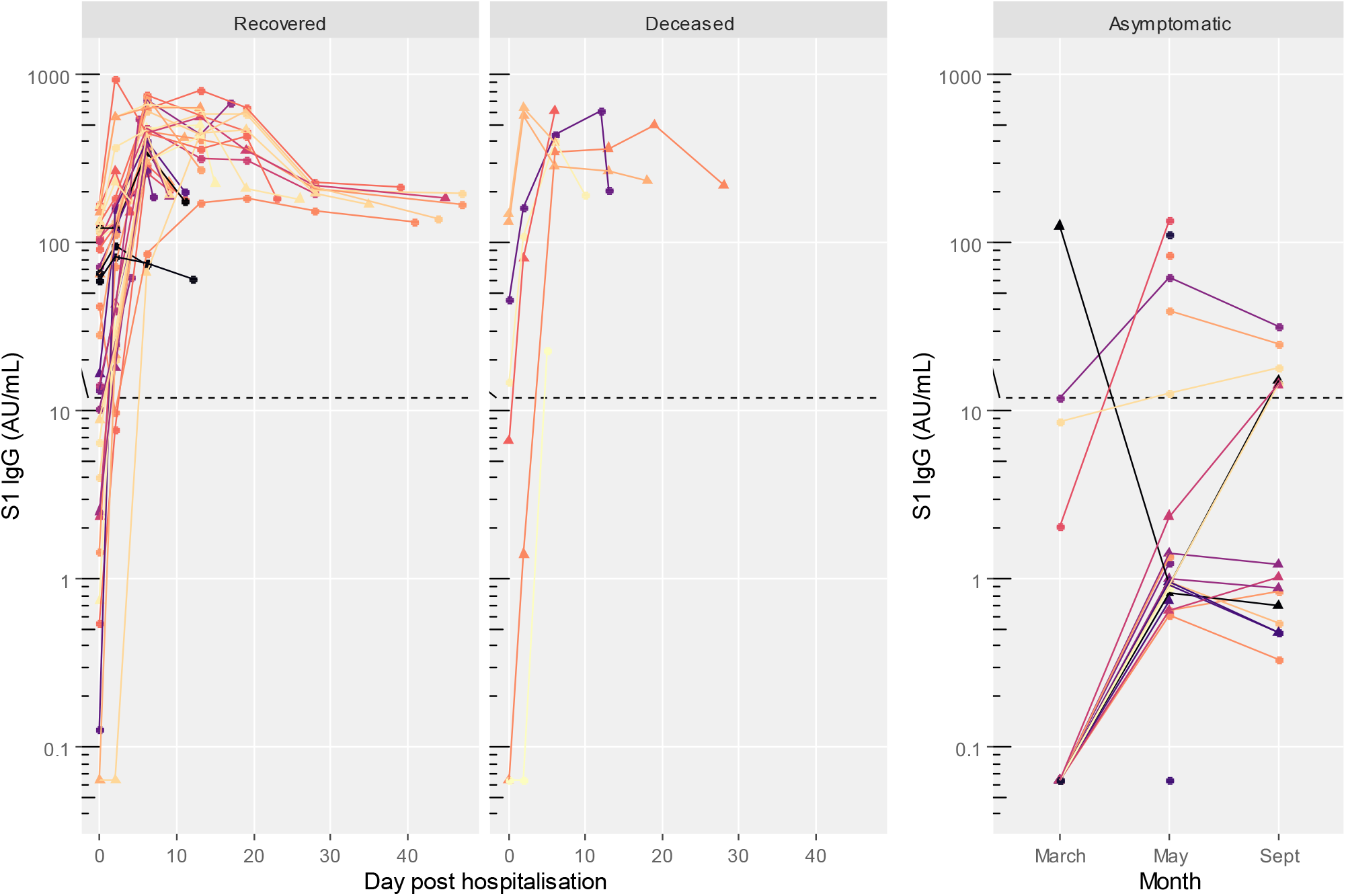
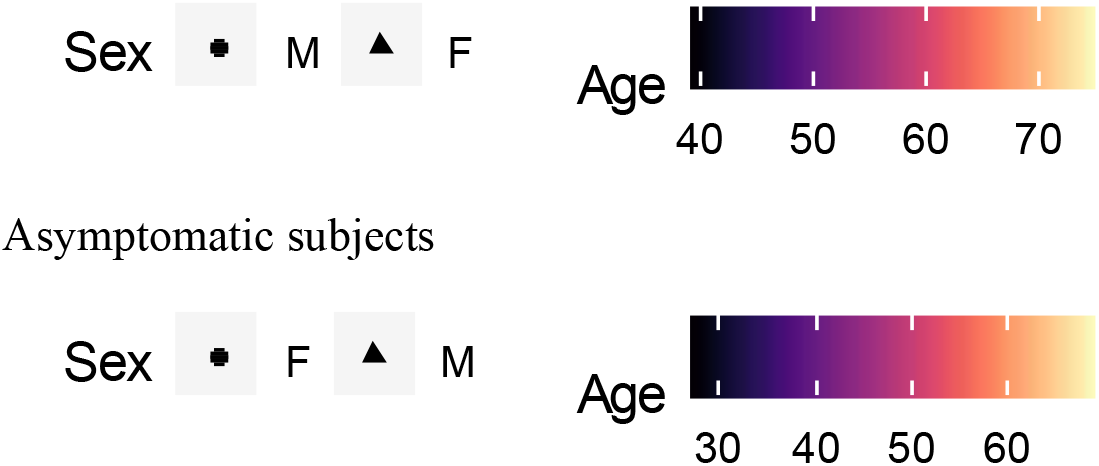
S1 IgG titres in COVID-19 patients (recovered and deceased) and asymptomatic subjects. Black dashed line indicates positivity threshold at 12 U/ml.

**Figure 4.**
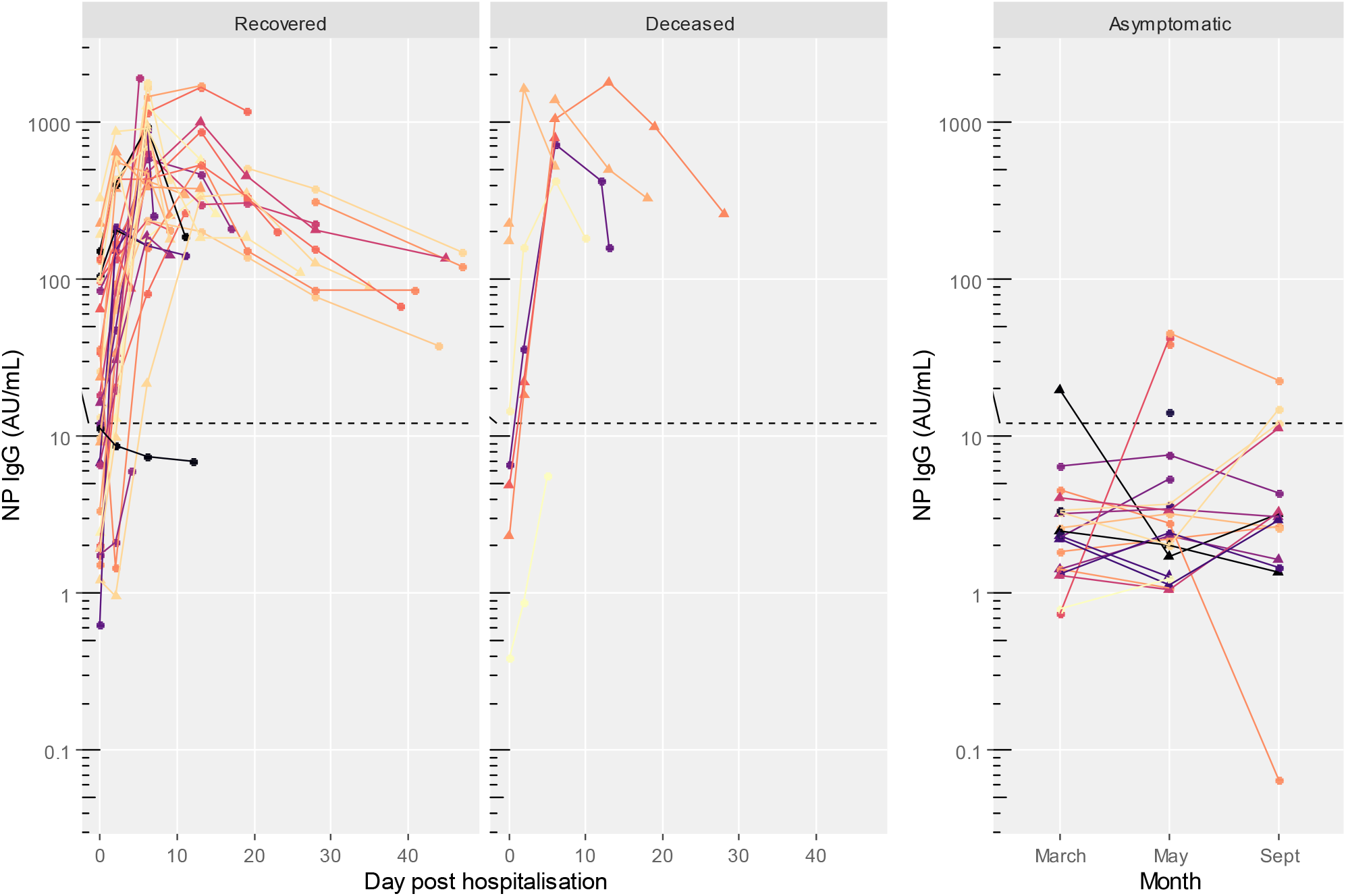
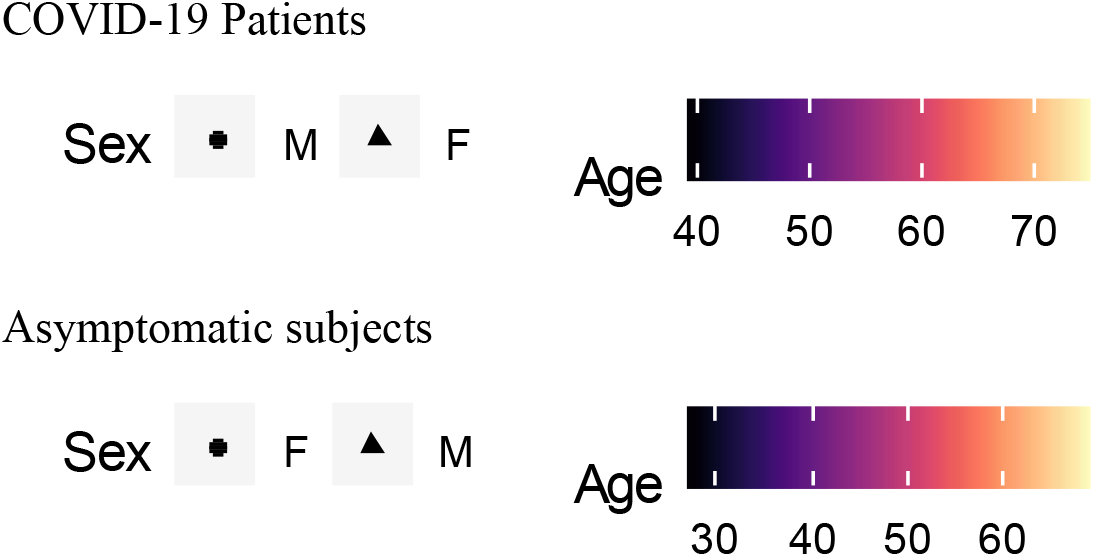
NP IgG titres in COVID-19 patients (recovered and deceased) and asymptomatic subjects. Black dashed line indicates positivity threshold at 12 U/ml.

**Figure 5.**
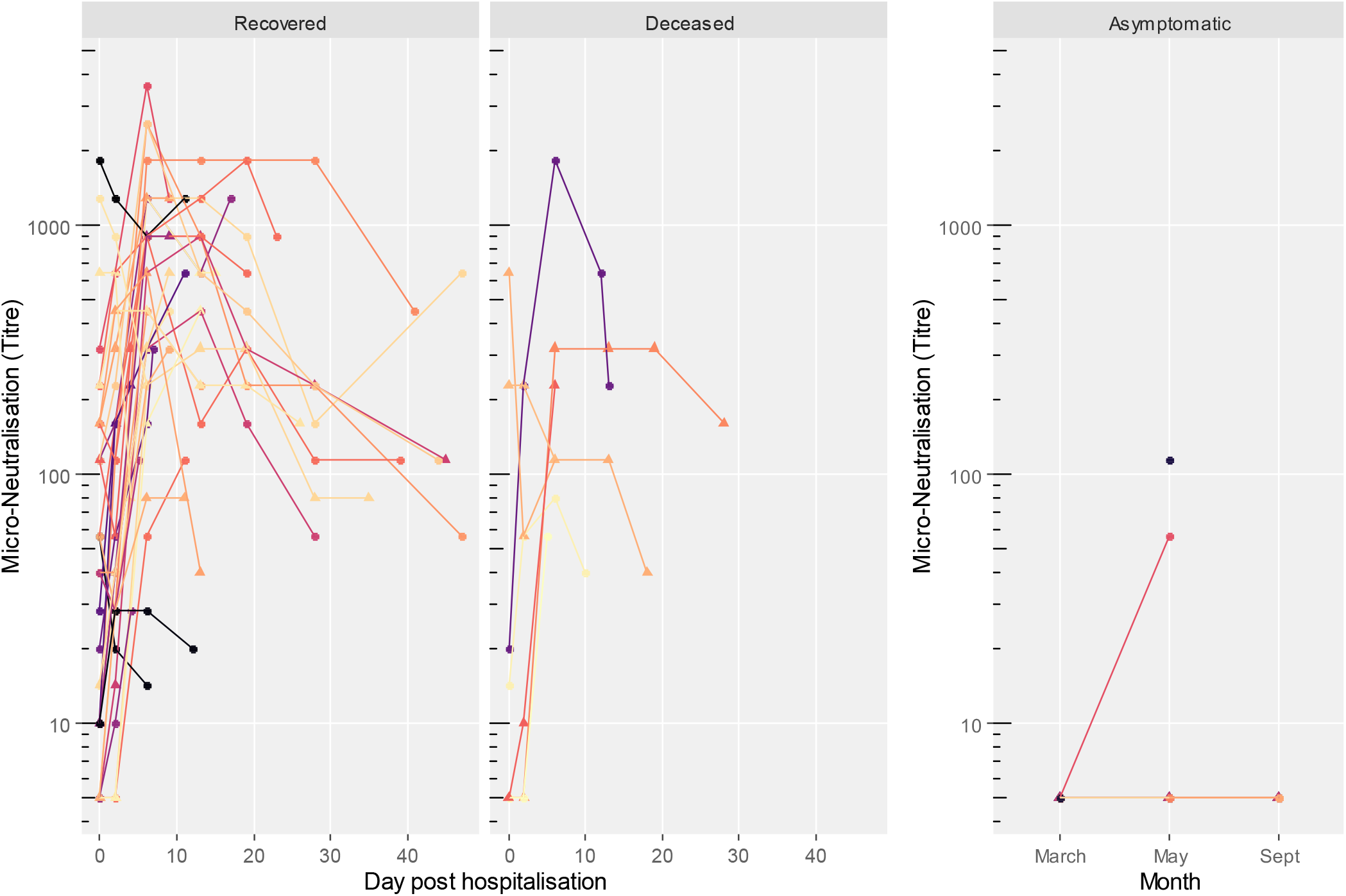
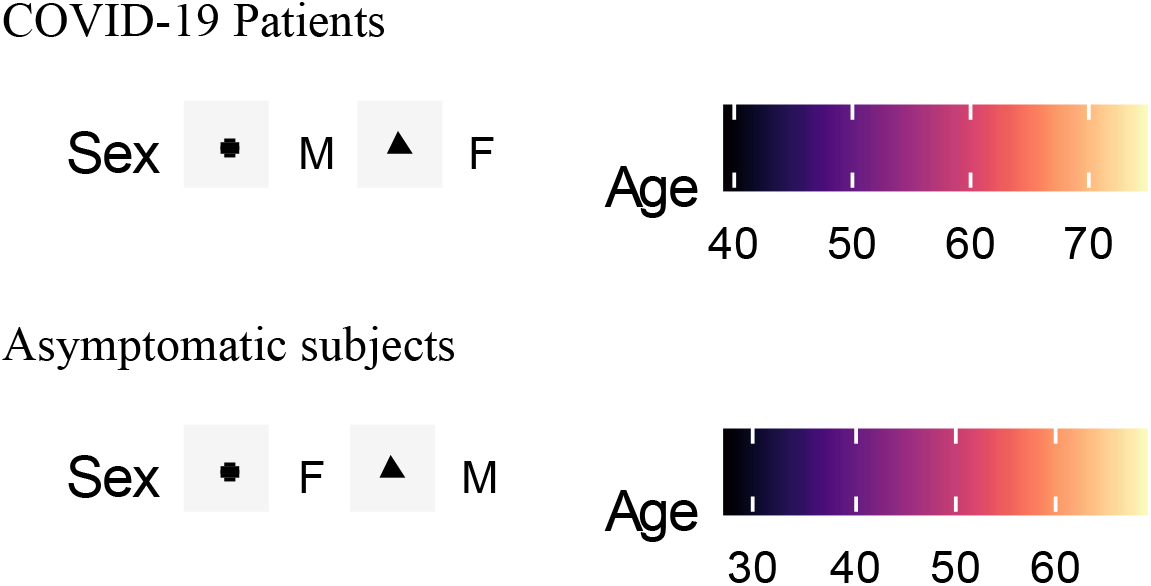
Neutralizing antibody (NAb) titres in COVID-19 patients (recovered and deceased) and asymptomatic subjects.

Neutralizing antibodies were found in all patients, with a range from 10 to 5120.

Antibody titers for patients are presented in table 2. S1 and NP antibodies started increasing at day 2 and again at day 6. A decrease for all antibodies was observed in recovered patients at day 27-30. S1 antibody increase was similar in both recovered and deceased patients, while NP IgG titers were significantly higher in deceased patients at day 6 (p-value 0.044). At baseline neutralizing antibody titers in recovered patients were 40.8 (95%CI 1.3 -1296.4) and 24.4 (95%CI 0.2-3093.8) in those deceased. Already at day 6, neutralizing antibody titers had increased steadily with 427.9 (95% CI 29.0 - 6321.5) in recovered patients and 226.3 (95% CI 12.1 -4228.2) in those deceased and showed a plateau in recovered patients until day 18-20. At day 27-30, neutralizing antibody titers had declined in recovered. No significant difference between the two groups, however comparison for the last days collection was not possible for few subjects in the deceased group.

**Table 2.**
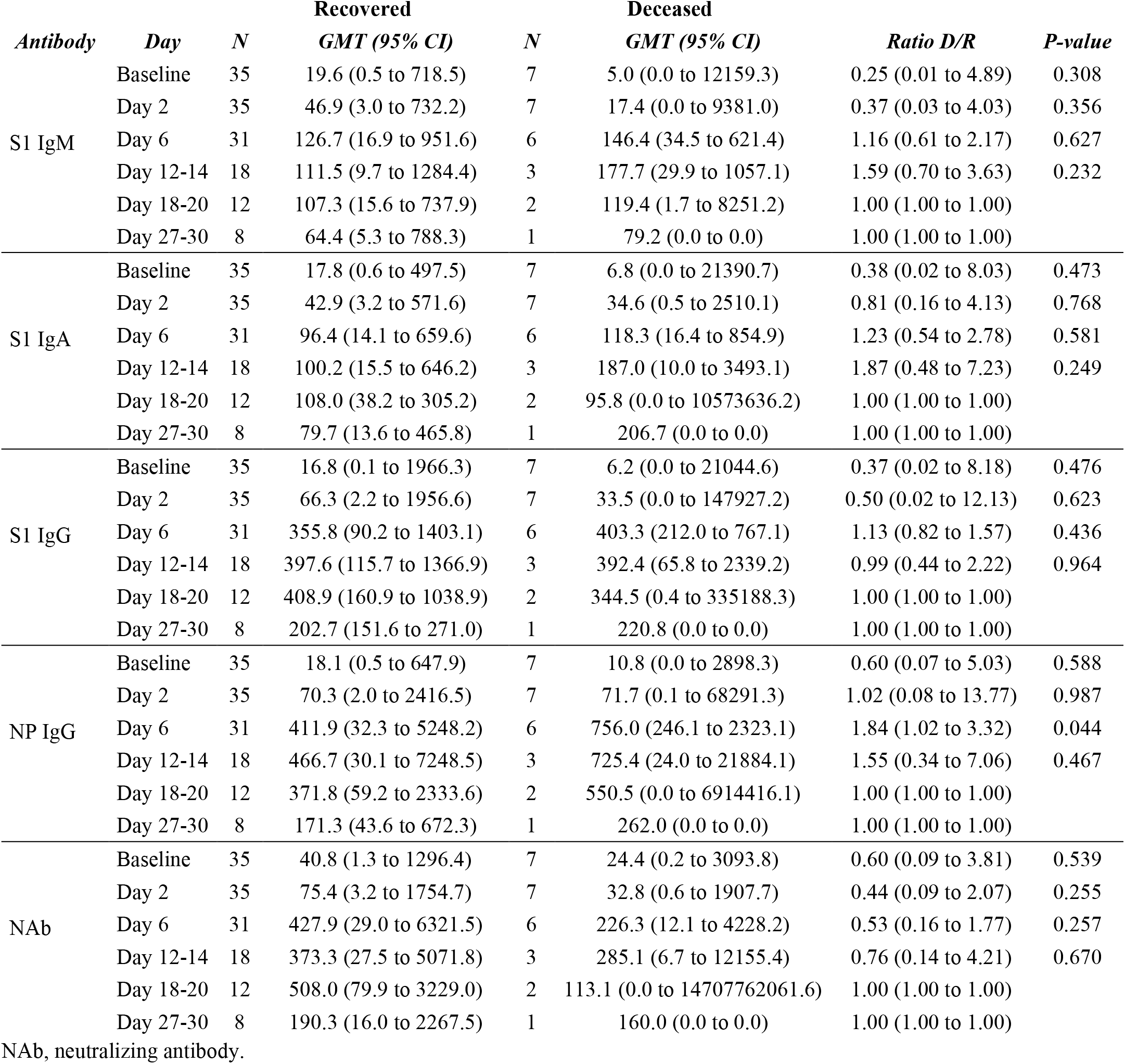
**Comparison of immune responses in recovered versus deceased COVID-19 patients. ELISA titres are expressed as U/ml**

Considering seroconversion rates in comparison to baseline (Table 3), IgM seroconversion rate appeared higher in the deceased (p-value 0.043).

**Table 3.** **Seroconversion in COVID-19 patients according to outcome. Seroconversion (SC) was calculated as 4-fold increase in titre compared to baseline**

| *Antibody* | *Day* | *Recovered* |  | *Deceased* |  | *P-value* |
| --- | --- | --- | --- | --- | --- | --- |
|  |  | *SC Yes* | *SC No* | *SC Yes* | *SC No* |  |
| S1 IgM | Day 2 | 6 | 29 | 4 | 3 | 0.043 |
|  | Day 6 | 19 | 12 | 4 | 2 | 1.000 |
|  | Day 12-14 | 11 | 7 | 2 | 1 | 1.000 |
|  | Day 18-20 | 7 | 5 | 1 | 1 | 1.000 |
|  | Day 27-30 | 4 | 4 | 1 | 0 | 1.000 |
| S1 IgA | Day 2 | 8 | 27 | 3 | 4 | 0.353 |
|  | Day 6 | 20 | 11 | 4 | 2 | 1.000 |
|  | Day 12-14 | 12 | 6 | 2 | 1 | 1.000 |
|  | Day 18-20 | 8 | 4 | 1 | 1 | 1.000 |
|  | Day 27-30 | 5 | 3 | 1 | 0 | 1.000 |
| S1 IgG | Day 2 | 11 | 24 | 5 | 2 | 0.085 |
|  | Day 6 | 23 | 8 | 4 | 2 | 0.653 |
|  | Day 12-14 | 13 | 5 | 2 | 1 | 1.000 |
|  | Day 18-20 | 9 | 3 | 1 | 1 | 0.505 |
|  | Day 27-30 | 4 | 4 | 1 | 0 | 1.000 |
| NP IgG | Day 2 | 16 | 19 | 6 | 1 | 0.096 |
|  | Day 6 | 24 | 7 | 5 | 1 | 1.000 |
|  | Day 12-14 | 14 | 4 | 2 | 1 | 1.000 |
|  | Day 18-20 | 9 | 3 | 1 | 1 | 0.505 |
|  | Day 27-30 | 7 | 1 | 1 | 0 | 1.000 |
| NAb | Day 2 | 5 | 30 | 1 | 6 | 1.000 |
|  | Day 6 | 22 | 9 | 4 | 2 | 1.000 |
|  | Day 12-14 | 13 | 5 | 2 | 1 | 1.000 |
|  | Day 18-20 | 9 | 3 | 1 | 1 | 0.505 |
|  | Day 27-30 | 6 | 2 | 1 | 0 | 1.000 |

NAb, neutralizing antibody.

No statistically significant difference in antibody titers at baseline and by peak antibody level (all 5 assays combined) was found between those survivors and deceased by using the Cox proportional hazard model. A good correlation was found among all assays (Figure 6). Overall, the level of S1 specific response was well correlated among antibody types (r=0.781 and r=0.794, S1 IgG correlating with S1 IgA and S1 IgM, respectively; r=0.760 S1 IgA correlating with S1 IgM). S1 IgG response was highly correlated with NP IgG (r=0.834). Neutralizing antibodies well correlated with all ELISA antibodies tested (r=0.722 with S1 IgA, r=0.798 with S1 IgM, r=0.739 with S1 IgG, and r=0.730 with NP IgG).

**Figure 6.**
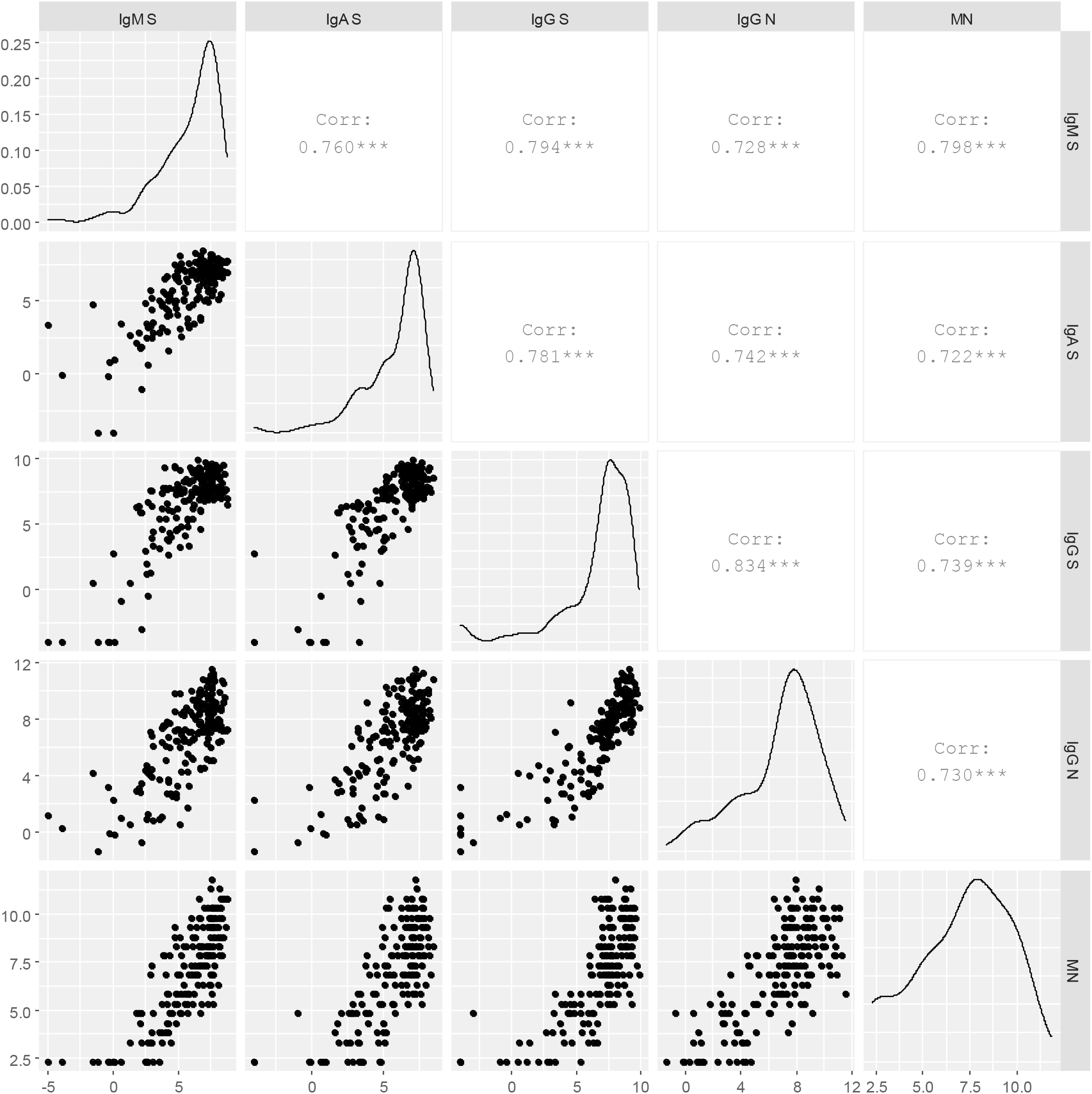
Correlations between antibody for COVID-19 patients. Titres are shown as log-2 transformed.

### Asymptomatic SARS-CoV-2 infected subjects

Asymptomatic infected subjects serum was made available as part of the UNICORN project [23]. The median age of asymptomatic subjects was 45.0 years (IQR 36.0-60.0), 8 were males and 17 females (Table 1b). Twenty-one subjects had the swab positive to SARS-CoV-2 in March 2020 and a blood draw, of whom 19 subjects had a blood draw in May and 14 another blood draw in September. Four subjects had the swab positive to SARS-CoV-2 in May and a blood draw, of whom 1 had a second blood draw in September.

**Table 1b.**
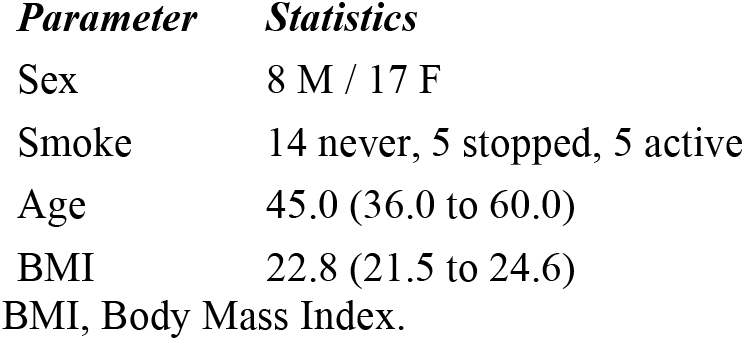
Baseline characteristics of SARS-CoV-2 asymptomatic subjects. Median (IQR)

Out of 25 asymptomatic subjects, 16 (64.0%) were negative to any antibody at any time point. Nine subjects (36.0%) had at least one detectable antibody type at least at one of the time points. At the first time point 6 subjects (24.0%) had positive S1 IgG, of whom 2 also had S1 IgA and NP IgG, 1 also S1 IgM, S1 IgA and NP IgG and 1 had S1 IgA, NP IgG and positive neutralizing antibodies. Three subjects negative at any antibody at the first time points had antibodies at one of the subsequent time points. One of these subjects was positive at any antibody assay at the second time point including neutralizing antibodies. The other 2 subjects had S1 IgA, S1 IgG and NP IgG at the third time point. One subject was positive to all ELISA antibodies (S1 IgM, S1 IgA, S1 IgG, and NP IgG) at the first time point, negative at the second time point, and positive again only to S1 IgG at the third time point. Detectable neutralizing antibodies were found only in 2 subjects (8.0%), one at the first and only time point available and in the other one at the second time point, the third time point wasn’t available. Both subjects were also positive to S1 IgA, S1 IgG, and NP IgG; only the first one was also positive to S1 IgM.

Asymptomatic subjects with positive antibody levels in any of the assays had titers well below the level found in symptomatic subjects as shown in figures 1-5 (table 4).

**Table 4.** Comparison of baseline immune responses for Recovered or Deceased versus asymptomatic controls.

| *Antibody* | *Recovered/ Asymptomatic* | *P value* | *Deceased / Asymptomatic* | *P value* |
| --- | --- | --- | --- | --- |
| S1 IgM | 286 (111 to 734) | 1.42E-15 | 73 (4 to 1448) | 0.0111 |
| S1 IgA | 172 (71 to 422) | 6.05E-15 | 65 (3 to 1351) | 0.0145 |
| S1 IgG | 96 (27 to 338) | 4.86e- 9 | 36 (2 to 798) | 0.0295 |
| NP IgG | 7.3 (3.7 to 14.3) | 3.22e- 7 | 4.3 (0.5 to 36.3) | 0.142 |
| NAb | 8.2 (4.6 to 14.6) | 1.88e- 8 | 4.9 (0.8 to 30.5) | 0.0785 |

## DISCUSSION

In this study we primarily evaluated the time course of the antibody response to different antigens of SARS-CoV-2 (IgG, IgM, and IgA against S1, IgG against NP, and neutralizing antibodies) in COVID-19 patients admitted to hospital for pneumonia during the first epidemic wave in March and April 2020 in Italy. No significant difference in titers was observed in any of the S1 antibody class at any time point between patients who survived and who did not survive. However, in deceased patients, a sharper increase of S1 antibodies has been observed suggesting a potential risk factor. Notably, IgM seroconversion rate appeared somewhat higher in the deceased, which might indicate that the deceased were admitted early after infection. In other similar studies early antibody response to S1 IgA or IgM or difference in the magnitude of the immune response to SARS-CoV-2 infection was a predictor of disease severity or progression or outcome [25-28]. IgG antibody titers against NP were significantly higher in the deceased at day 6 according to other studies in which an early response to NP in the first 15 days post-disease onset was indicative of fatal outcomes [25, 27]. Our results did not highlight any difference for neutralizing antibodies in contrast with other studies in which these antibodies were significantly higher in patients who required ICU or died [27]. One explanation for the similar immune response between the two study groups can be that in our study COVID-19 patients had, at admission, similar clinical and demographic characteristics. However, other factors could be implicated in the specific immune response against SARS-CoV-2 that need to be further investigated, as other elements of cellular-mediated immunity may play a role in protection. The kinetics of the antibody response showed an increase starting from day 2 and reaching the peak between days 6 and 18-20. At 27-30 day, a decline in titers was observed for any of the antibody classes. In other studies, the antibody kinetic in COVID-19 patients showed the antibody peak reaching up to the 4^th^ and 5^th^ week from disease onset followed by antibody decay starting at the 6^th^ week [28, 29]. This observation differs from our findings most likely as our study period of patients started at hospital admission when the severity of the disease was already in an advanced stage.

In this study, a high correlation between S1 and NP protein-based ELISAs and the neutralization assay was observed in COVID-19 patients. The reason for that is likely due to the fact that the S1 protein represents the immunodominant antigen of SARS-CoV-2 virus and functional neutralizing antibodies are mainly targeting the Receptor-Binding Domain, a subunit of that protein [30].

In this study, only few asymptomatic subjects had detectable titers with almost two-thirds negative at any time point. These findings support the observation that the use of serology testing for population surveys might account of false negative results as asymptomatic or paucisymptomatic subjects might have antibody concentration below the level of detection [31]. Only few asymptomatic subjects had detectable titers to S1 IgG or more antibodies but with a low level of titers as compared to COVID-19 patients included in this study in accordance with another study [32]. Notably, in contrast to COVID-19 patients, the majority of asymptomatic subjects do not have circulating neutralizing antibodies that are considered a surrogate of protection against COVID-19, thus vaccination is highly recommended. Although further evidence should be provided to establish a correlate of protection, based on these data it can be speculate that evidence of neutralizing antibodies might be used as markers of protective immunity.

Asymptomatic subjects have a low viral load in the nasopharynges as assessed by RT-PCR and most likely a lack or a defective viral replication and/or mucosal invasion that induces a weak or any antibody response [33]. Although memory B and T cells may have been primed in SARS-CoV-2 swab positive subjects with undetectable antibodies in serum, with the ability to induce a rapid immune response to re-exposure, the question of whether these subjects should be vaccinated is critical now that effective vaccines are available against COVID-19.

One limitation of this study is its retrospective nature and the collection of COVID-19 samples was carried out only in a single center. Besides this, we limited the collection to those patients for whom it was possible to construct an antibody response curve over a period of at least one month. This, of course, represents a bias, however, the ratio between deceased and recovered patients (7 out of 42), 16.6% falls in the range from 5.7% to 30.4% as described in the literature [34, 35]. This value shows high variability because it can be influenced both by the characteristics of the series studied and the different treatments. The findings from this study do not allow us to predict the kinetics of the antibody decay over time in patients who recovered from COVID-19, in particular, who will be susceptible to reinfection over time, since no follow-up samples after discharge were available.

Overall, our data highlight that COVID-19 patients produce a simultaneous antibody response to SARS-CoV-2 with high correlation between different viral antigens (S1 and NP) and among antibody classes (IgA, IgG, and IgM and neutralizing antibodies). The peak is reached by 3 weeks from hospital admission followed by a sharp decrease. On the contrary, only few asymptomatic subjects developed antibodies at detectable levels, though significantly lower compared to COVID-19 patients. Since neutralizing antibodies were rarely produced, this finding raises the question about the protection of these subjects against reinfection.

## Data Availability

Data sharing not applicable. No new data were created or analyzed in this study.

## Author’s contributions

S.M.: investigation, data curation, writing – review and editing; S.V.: conceptualization, funding acquisition, writing – original draft preparation; E.J.R.: formal analysis, writing – review & editing; A.R.: resources, data curation, writing – review & editing; E.B.: resources, data curation, writing – review & editing; V.B.: resources, data curation, writing – review & editing; G.P.M: resources, data curation, writing – review & editing; A.M.: methodology, writing – review and editing; G.L.: methodology, writing – review and editing; A.R.; methodology, writing – review and editing; E.M.: methodology, writing – review and editing; C.M.T.: conceptualization, funding acquisition, investigation, project administration, writing – review and editing.

## Acknowledgements

This study was funded by a research grant (Pfizer Tracking Number 60353289).

This funding source did not have any role in the design of this study and during its execution, analyses, interpretation of the data, or decision to submit results.

This study was supported by the European Virus Archive goes Global (EVAg) project, which has received funding from the European Union’s Horizon 2020 research and innovation programme under grant agreement No 653316.

The UNICORN population was recruited thanks to the Funding Action “Ricerche Emergenza coronavirus”, University of Milan, 2020.

The authors thank Linda Benincasa for technical support.

